# Confirmed and Unreported COVID-19-Like Illness Death Counts: An Assessment of Reporting Discrepancy

**DOI:** 10.1101/2020.07.20.20158139

**Authors:** Mazbahul G. Ahamad, Fahian Tanin, Byomkesh Talukder

## Abstract

**Objective:** To assess the reporting discrepancy between officially confirmed COVID-19 death counts and unreported COVID-19-like illness (CLI) death counts.

**Study Design:** The study is based on secondary time-series data.

**Methods:** We used publicly available data to explore the differences between confirmed COVID-19 death counts and deaths with probable COVID-19 symptoms in Bangladesh between March 8, 2020, and July 18, 2020. Both tabular analysis and statistical tests were performed.

**Results:** During the week ending May 9, 2020, the unreported CLI death count was higher than the confirmed COVID-19 death count; however, it was lower in the following weeks. On average, unreported CLI death counts were almost equal to the confirmed COVID-19 death counts during the study period. However, the reporting authority neither considers CLI deaths nor adjusts for potential seasonal influenza-like illness or other related deaths, which might produce incomplete and unreliable COVID-19 data and respective mortality rates.

**Conclusions:** Deaths with probable COVID-19 symptoms needs to be included in provisional death counts in order to estimate an accurate COVID-19 mortality rate and to offer data-driven pandemic response strategies. An urgent initiative is needed to prepare a comprehensive guideline for reporting COVID-19 deaths.

## 1. Introduction

Underreporting COVID-19 death statistics, either by mistake or deliberate, is not uncommon across the world, with countries including China and Italy adjusting their death counts.^1^ The World Health Organization (WHO) suggests including “death resulting from a clinically compatible illness in a probable or confirmed COVID-19 case”^2^ for pandemic surveillance. However, since the beginning of the pandemic, evidence has emerged showing the Institute of Epidemiology Disease Control and Research (IEDCR), a government-affiliated reporting institute in Bangladesh, has reported COVID-19 deaths without maintaining any standard reporting guidelines (e.g., WHO).

Between March 8 and July 18, 2020, a total of 2,580 COVID-19 deaths were officially recorded by the IEDCR.^3^ During the same period, a total of 1,874 people died with symptoms consistent with COVID-19-like illness (CLI), according to the Centre for Genocide Studies (CGS) in Bangladesh.^4^ That is 1,874 more deaths that were potentially caused by COVID-19 than were reported, which raises doubts about the reliability of existing COVID-19 death counts. On average, unreported CLI death counts were similar to officially confirmed COVID-19 death counts during the study period, which suggests a major data reporting discrepancy.

COVID-19 death counts and mortality rate have direct effect on research and policy implications as well as pandemic response strategies. Documentation of both confirmed and unreported death counts by the reporting authority has become essential. We, therefore, compared officially confirmed COVID-19 and unreported CLI death counts to explore any reporting discrepancy in the absence of a practical COVID-19 death reporting guideline.

## 2. Data and Findings

We used publicly available data to explore trends and assess the differences between confirmed COVID-19 and unreported CLI death counts between March 8, 2020, and July 18, 2020, in Bangladesh. Officially confirmed daily COVID-19 death data were obtained from www.worldometers.info. Unreported CLI death counts were collected from the weekly report of the CGS.^4^ The CGS collects COVID-19-like death counts from daily newspapers, retraces each case, and removes duplication. We calculated the weekly confirmed COVID-19 death counts to be comparable to weekly unreported CLI death counts. We also estimated the two-week moving average of both death counts. Both tabular and statistical analyses were conducted using Stata version 16.1.^5^ Data used in the study will be available publicly at Harvard Dataverse.^6^

Fig. 1 shows weekly counts of both confirmed COVID-19 and unreported CLI deaths. The mean number of confirmed deaths was 136 (95% CI: 76.60-194.98) per week, and the mean number of unreported CLI deaths was approximately 99 (95% CI: 65.55-131.72) per week. No statistically significant mean differences existed between confirmed COVID-19 and unreported CLI death counts (*p* = 0.257).

**Fig. 1.**
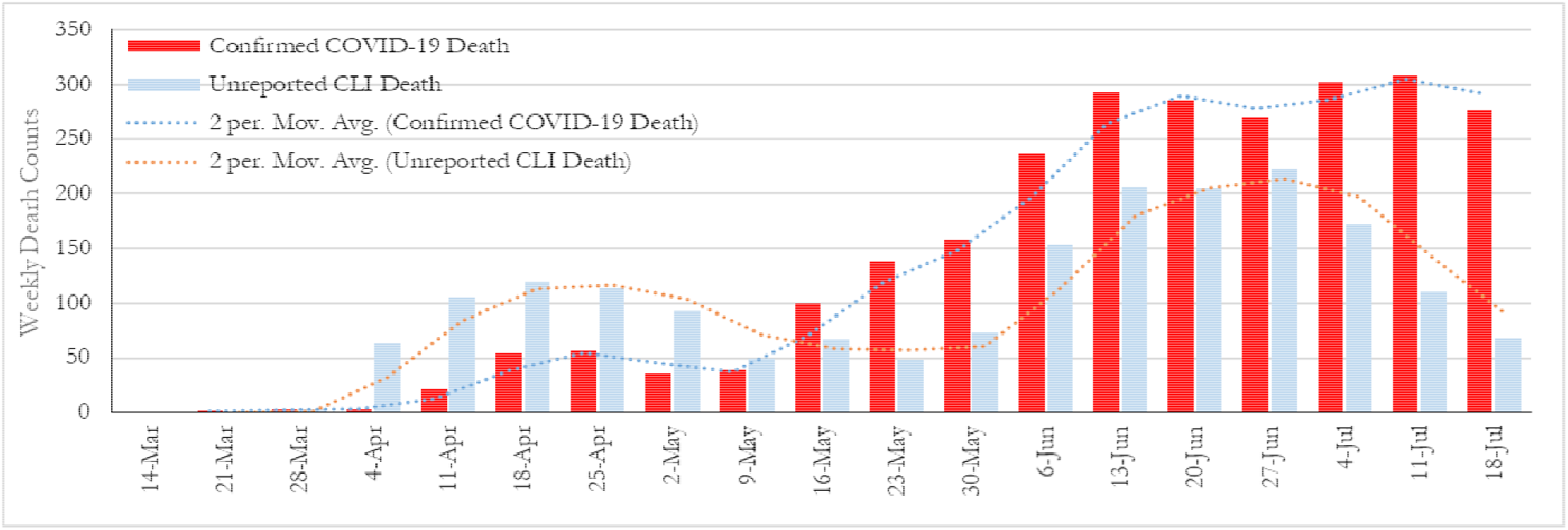
Weekly confirmed COVID-19 and unreported CLI death counts, March 8 — July 18, 2020, Bangladesh. The horizontal axis indicates the last date of the respective week.

Though both confirmed and unreported death counts show increasing trends over the study period in majority of the weeks, the unreported death counts exhibit a decreasing trend in recent weeks. For the week ending May 9, 2020, the confirmed COVID-19 death counts exceeded the unreported CLI death counts (Fig. 1). Before this week, unreported CLI death counts were higher than confirmed COVID-19 death counts (average: ∼61 vs 24); however, it was lower in the following weeks (average: ∼133 vs ∼237) (Table 1). On average, the percentage of unreported CLI deaths was 55% of total deaths (sum of confirmed and unreported) from March 8 to May 9, 2020, decreasing to 35% from May 10 to July 18, 2020.

**Table 1.**
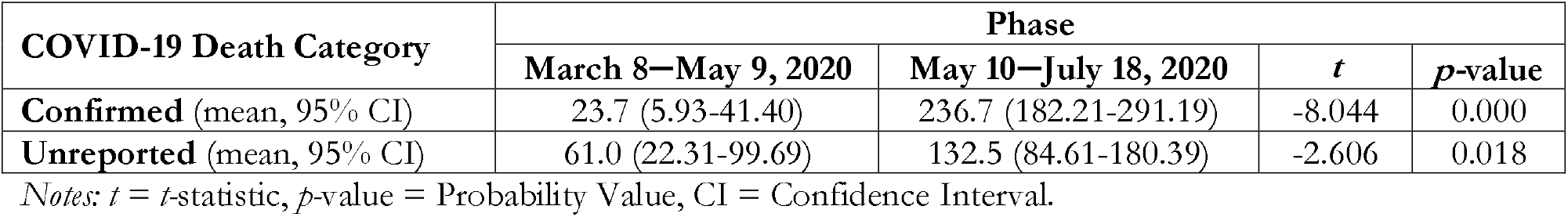
Mean confirmed COVID-19 and unreported CLI death counts, March 8 — July 18, 2020, Bangladesh.

| COVID-19 Death Category | Phase |  |  |  |
| --- | --- | --- | --- | --- |
|  | March 8–May 9, 2020 | May 10–July 18, 2020 | *t* | *p*-value |
| **Confirmed** (mean, 95% CI) | 23.7 (5.93-41.40) | 236.7 (182.21-291.19) | -8.044 | 0.000 |
| **Unreported** (mean, 95% CI) | 61.0 (22.31-99.69) | 132.5 (84.61-180.39) | -2.606 | 0.018 |

Notes: *t* = *t*-statistic, *p*-value = Probability Value, CI = Confidence Interval.

## 3. Discussion

The percentage of CLI deaths that were unreported is high; almost half (44%) of the total deceased who showed COVID-19-like symptoms were either not tested after death or not reported appropriately. The reporting authority neither documented these deaths as resulting from COVID-19-like illness nor adjusted for potential seasonal influenza-like illness and other related deaths per the relevant guidelines,^7,8^ which might produce unclear COVID-19 data and mortality rates. This significant number of unreported CLI deaths may incorrectly suggest that the prevalence of COVID-19 in Bangladesh is low; this, in turn, can be problematic from two different, but interlinked perspectives.

First, public health researchers and policy analysts will not properly recognize the dynamics of COVID-19 without reliable data. This will restrict their ability to accurately identify the current pandemic stage of COVID-19 and predict the peak case and death counts. Second, the death reporting gap may present challenges when preparing COVID-19 response strategies, for example, in allocating coronavirus treatment hospitals and intensive care beds. Incorrect death counts can undermine ongoing response efforts and limit access to public health assistance, such as vaccines, coming from international organizations and development partners.

In conclusion, we recommend that the IEDCR responsibly implement real-time tracking and reporting of CLI deaths as provisional along with confirmed COVID-19 deaths, complying with the World Health Organization’s suggested definition of COVID-19 deaths,^2^ which could be similar to the reporting standard of the Centers for Disease Control and Prevention of the United States.^9^ Both confirmed and provisional death statistics will enable public health researchers, policy analysts, and development partners to correctly estimate the COVID-19-related mortality rate and offer data-driven pandemic preparedness, early responses, and emergency actions with necessary national and international public health assistances.

## Data Availability

Supplementary data and code of the article can be found at Harvard Dataverse (https://doi.org/10.7910/DVN/H9RHWN) upon request.

https://dataverse.harvard.edu/dataset.xhtml?persistentId=doi:10.7910/DVN/H9RHWN

## Author statements

### Acknowledgement

We thank the Centre for Genocide Studies (www.cgsdu.org) for collecting and disseminating data regarding death counts with COVID-19-like symptoms.

### Author’s Contributions

MGA conceived the original idea and gathered data jointly with FT. MGA analyzed the data and wrote the first draft using literature jointly sourced by MGA, FT, and BT. MGA and BT edited several drafts to produce finally approved manuscript. MGA has been published an <u>opinion article</u> based on this manuscript in *The Business Standard* on June 2, 2020.

### Ethical approval

Ethical approval was not sought as this study used publicly available data.

